# The Role of Pain Frequency in the Divergent Associations Between Cannabis Use and Default Mode Network Connectivity

**DOI:** 10.64898/2026.03.10.26348038

**Authors:** Tracy Brown, Che Liu, Emese Kroon, Janna Cousijn, Francesca Filbey

**Affiliations:** Department of Psychology, University of Texas at Dallas, Dallas, TX, USA; Erasmus University, Rotterdam, The Netherlands

**Keywords:** fMRI, cannabis, pain, DMN, rsFC

## Abstract

**Background:** Chronic pain is one of the most common reasons for medicinal cannabis use, yet the neural mechanisms underlying cannabis-related modulation of pain remain poorly understood. Both pain and cannabis use independently alter functional connectivity within the brain’s default mode network (DMN) that modulate interoception and self-referential aspects of pain processing. The goal of this study was to examine the interaction between pain and cannabis use on DMN connectivity.

**Methods:** We measured DMN resting state fMRI functional connectivity (rsFC), past year pain frequency, and cannabis use measures (i.e., grams per day, days a week, years of regular use) from 119 adults who use cannabis near-daily (68 men; Mage= 22.66, SE= .31). Generalized linear models were used to test the main effects and interactions of pain frequency and cannabis use variables.

**Results:** Results indicated significant interactions between pain and cannabis use where more frequent pain was (1) negatively associated with weekly use or years of use in l-IPL-PCC and r-IPL-PCC rsFC, (2) whereas it was positively associated with daily grams of cannabis in l-IPL-r-IPL rsFC and r-IPL-PCC rsFC (BH-FDR-corrected p< .05).

**Conclusions:** First, these findings demonstrate that pain frequency is a key context shaping the neurobiological correlates of exposure to cannabis. Second, divergent interaction effects suggest that, in the context of more frequent pain, cannabis use may relate to rsFC through distinct neural processes that depend on cumulative vs. proximal effects.

## Introduction

Chronic pain is one of the most common reasons for medicinal cannabis use (1,2). Those who use cannabis to alleviate pain symptoms show greater escalation of use over time compared to those who use cannabis for other symptoms, such as insomnia, depression, or anxiety (1). This escalation of cannabis use among those with chronic pain confers an elevated risk for developing a cannabis use disorder (CUD) (2). Given the high prevalence of medicinal use, the propensity for increasing use, and the heightened vulnerability for CUD in those with chronic pain, there is a critical need to examine how pain-related cannabis use influences pain-regulating brain networks.

The sensory, affective, and cognitive dimensions of pain are regulated across the primary and secondary somatosensory cortices, insula, cingulate cortex, thalamus, amygdala, orbitofrontal cortex, and prefrontal regions (3–5). The brain exerts top-down control of pain via descending analgesic systems that involve the periaqueductal gray (PAG) nucleus raphe magnus (NRM), which inhibits pain transmission in the spinal dorsal horn (6). These cortical and subcortical pain-modulating brain regions are functionally integrated within resting-state brain networks (RSNs) such as the default mode network (DMN), which plays a key role in mediating nociceptive processing and subjective pain experience (7).

Chronic pain has been reported to weaken rsFC between the posterior cingulate cortex (PCC) and medial prefrontal cortex (mPFC) as well as between PCC and inferior parietal lobes (IPL) (8,9). This reduction in rsFC has been related to maladaptive self-referential processing, rumination about pain, and difficulties disengaging attention from pain (10). Reduced DMN rsFC also relates to clinical measures of chronic pain such as higher subjective pain ratings, reporting more frequent pain, and more years of experiencing pain (11,12). These findings suggest that chronic pain contributes to an intricate dysregulation of top-down pain-modulatory systems, which influence the degree of perceived pain (9,10). Similarly, cannabis use has been related to alterations to DMN rsFC that may moderate pain signaling. Studies have shown that frequency (>40 past year uses) and duration of cannabis use (i.e., 500+ lifetime uses) reduce DMN rsFC (13–16). Neuroimaging studies show that chronic cannabis use reduces CB1 receptor availability within DMN regions (17,18). CB1 receptors regulate presynaptic release of glutamate and GABA and are critical for maintaining the excitation-inhibition synchrony (19). Alterations to these neuromodulatory systems may indirectly influence DMN coherence by shifting the balance between internally and externally oriented cognitive states. Over time, such shifts may promote DMN alterations characterized by weaker integration between posterior regions and resemble connectivity patterns that are similar to those with chronic pain (9). Thus, it is possible that cannabis may worsen chronic pain through the dysregulation of top-down pain regulatory processes via decreased DMN connectivity.

Our review of the literature suggests that ongoing pain and cannabis use independently reduce rsFC between DMN regions that are implicated in pain regulatory processes, but how they potentially interact has not yet been examined. This study addressed this gap by examining whether the experience of pain and cannabis use interact to predict rsFC within the DMN. We hypothesized that pain would moderate the relationship between cannabis use on DMN rsFC, such that greater pain frequency and heavy, prolonged cannabis use would be associated with greater DMN rsFC reductions.

## Methods

The Institutional Review Board of the University of Texas at Dallas and the Department of Psychology of the University of Amsterdam ethics committee approved this study.

### Participants

Participants were recruited for a study that examined the neurocognitive effects of cannabis in a cross-cultural sample of adults (20–22). A total of 133 adults who use cannabis near-daily were from two study sites located in Dallas, Texas (*n* = 75) and the Netherlands (*n* = 59).

### Behavioral Measures

Pain frequency was assessed using a single item from the Self-rated Level 1 cross-cutting Symptom Measure (23) that asked, “During the past year, how much (or how often) have you been bothered by unexplained aches and pains (e.g., head, back, joints, abdomen, legs)?”. Participants answered using the following responses: never, sometime, half the time, most of the time, or all of the time. Cannabis use was measured as grams used per day, days a week of use, and years of weekly use, which were derived from the Substance Use History Questionnaire (SUHQ) (24).

### Resting State fMRI (rsfMRI) Scan Acquisition and Processing

High-resolution structural MRI scans were collected using the following parameters: T1 fast field echo; TR = 8.3 s, TE = 3.9 ms, 220 slices, 1 mm slice thickness, FOV = 240 × 188 × 220 mm, 1 × 1 mm voxel size, and a flip angle of 8°. rsfMRI scans were collected with eyes closed using T2* single-shot multiband-accelerated EPI sequence with the following parameters: TR = 0.55 s, TE = 30 ms, with 36 slices, 3 mm slice thickness and a 0.3 mm gap, FOV = 240 × 240 × 118.5 mm, 3 × 3 mm voxel size, and a flip angle of 55°.

Region-of-interest (ROI)-to-ROI connectivity values were extracted based on predefined atlas parcellations available within CONN for four DMN regions: mPFC (0, 52, -2), PCC (MNI: 0, -52, 26), l-IPL (-50, -63, 32), and the r-IPL (50, -63, 32). Functional connectivity between DMN regions was computed using Fisher Z-transformed bivariate correlation coefficients, and connectivity strength was indexed via beta weights derived from first-level general linear models (GLMs) using the CONN toolbox (25). This resulted in 6 rsFC scores as outcome measures.

### Statistical Analysis

Generalized linear models (GLM) are appropriate for modeling continuous neuroimaging outcomes and allow flexible specification of interactions between categorical and continuous predictors (25). We computed GLMs using likelihood ratio tests and Type III Wald χ² statistics to evaluate the significance of main effects and the interaction terms of pain frequency and cannabis use measures. To address imbalances across categorical pain frequencies and to provide stable parameter estimations, those reporting pain “half the time”, “most of the time”, or “all of the time” were collapsed into a single group that is consistent with standardized categorical indices of pain frequency (i.e., high vs. low) (26). Thus, pain frequency was entered as a three-level categorical factor (i.e., none: *n*=49; sometimes: *n*= 48; ≥ half the time: *n*=22). Cannabis use behaviors were included as continuous predictors given evidence that dose, frequency, and duration of cannabis exposure are associated with DMN connectivity (19,21). The interaction term between pain frequency and cannabis use behaviors was modeled to determine whether there were any cannabis-related alterations to rsFC strength and if these alterations were amplified from higher levels of pain. Age, gender, and study site (i.e., Netherlands vs. United States) were included as covariates of no interest.

To reduce Type I error, main effects and interactions were deemed significant if: (1) omnibus likelihood ratio tests indicated our model significantly predicted rsFC strength better than chance (*p*< .05), (2) overall predictor Wald statistic was significant (*p*<.05), (3) Type III effect *p*-values survived Benjamini-Hochberg (BH) FDR correction, and (4) conditional parameter estimates did not contain zero within the 95% confidence interval (27). We limited the number of models by including all cannabis use variables in each rsFC-strength model, thereby reducing the number of statistical tests and resulting in 6 GLMs.

The main effects of cannabis use behaviors (i.e., daily grams used, days of use per week, and years of weekly cannabis use) represent the association between cannabis use and rsFC strength within solely the “no pain” frequency. Moreover, the “no pain” frequency served as the reference slope for assessing the main effects of pain frequency and represented as the differences between “no pain” rsFC strength and the “sometimes” and “≥ half the time” pain frequencies, independent of the effects of cannabis. These effects are represented by the differences of rsFC strength. Lastly, the interaction coefficients indicated how much the cannabis-rsFC associations within the “sometimes” or “≥ half the time” pain frequencies differed from cannabis-rsFC associations with “no pain” (i.e., “no pain” main effects of cannabis).

## Results

### Study participants

Fourteen participants were excluded from the models due to missing cannabis use metrics, resulting in 119 individuals (68 men; Mage= 22.66, SE= .31) (see Table 1 for sample characteristics). Outliers, deemed as DMN ROI-ROI rsFC strength values ±3 SD from the grand mean, were excluded as they could bias the results (28) (see Table 2 for model inclusions). Omnibus likelihood ratio tests indicated that all but the mPFC-l-IPL rsFC model (χ²(14)= 21.18, *p*= .1) predicted DMN rsFC strength better than chance. Age, sex, and study site were not significantly associated with rsFC strength in any model (all *p* > .05) (see Table 3 for all results).

**Table 1.**
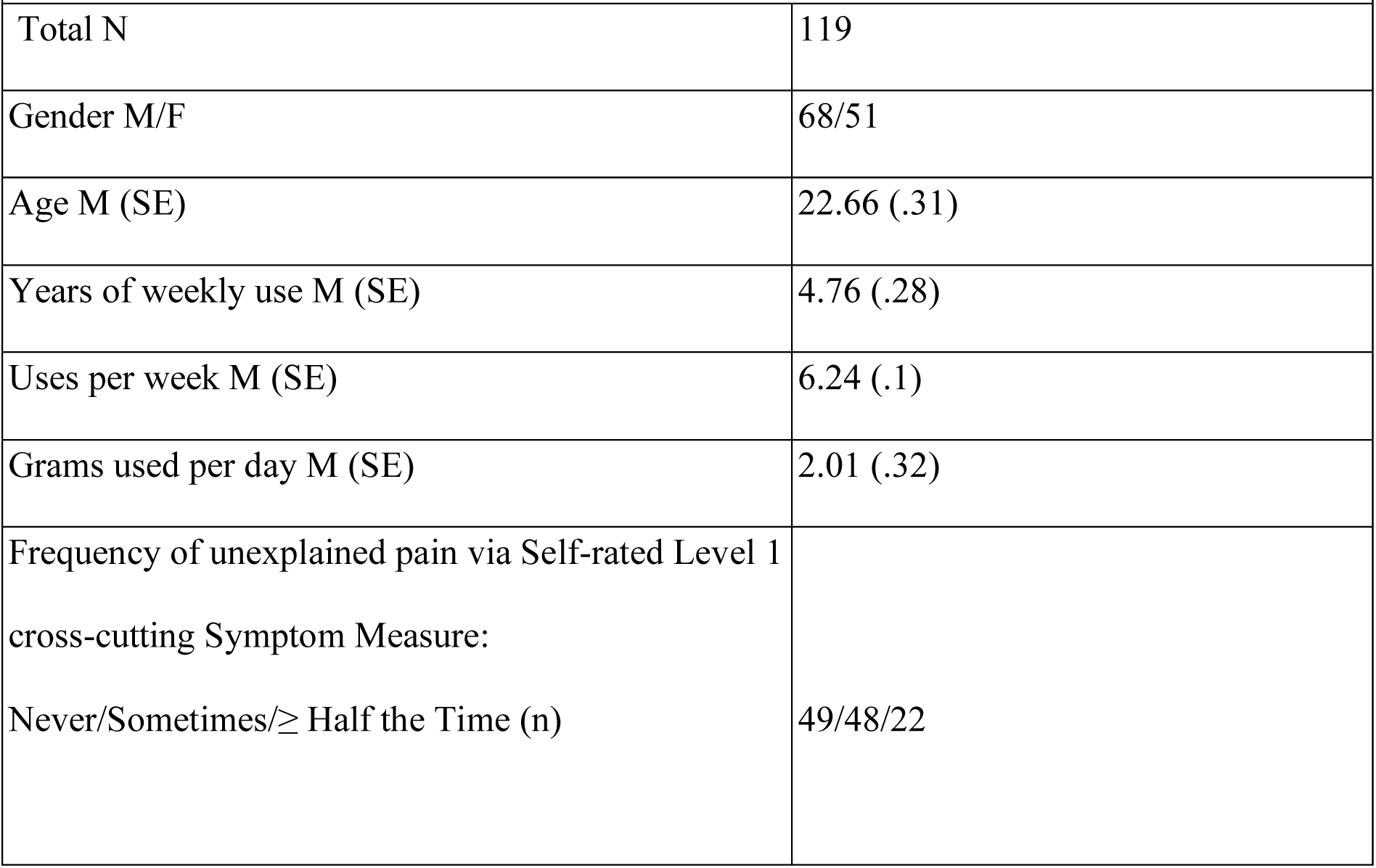
Sample Characteristics.

|  |  |
| --- | --- |
| **Table 1.** |  |
| *Sample Characteristics* |  |
| Total N | 119 |
| Gender M/F | 68/51 |
| Age M (SE) | 22.66 (.31) |
| Years of weekly use M (SE) | 4.76 (.28) |
| Uses per week M (SE) | 6.24 (.1) |
| Grams used per day M (SE) | 2.01 (.32) |
| Frequency of unexplained pain via Self-rated Level 1<br>cross-cutting Symptom Measure:<br>Never/Sometimes/≥ Half the Time (n) | 49/48/22 |

US= United States, NL= Netherlands

**Table 2.**
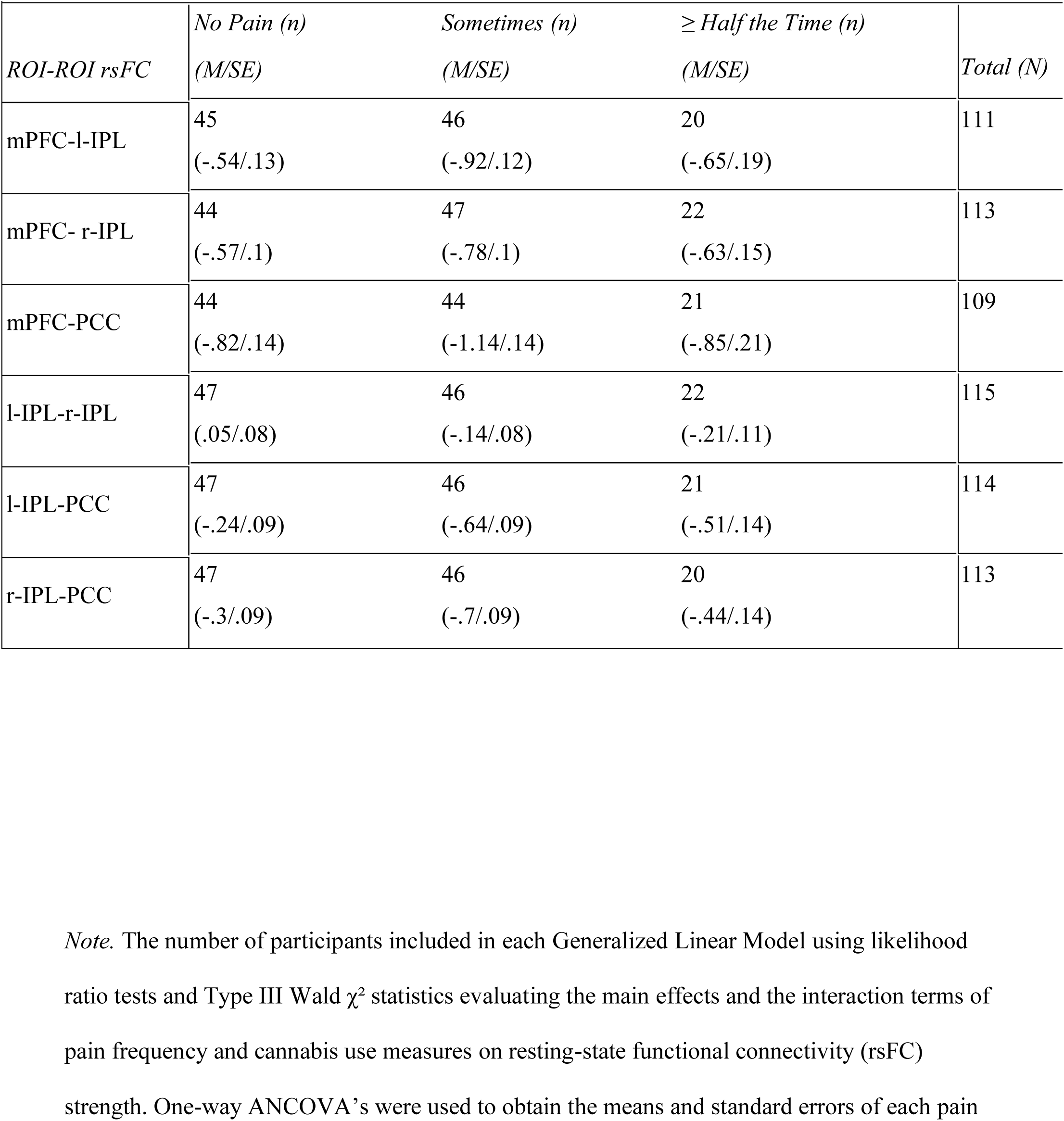
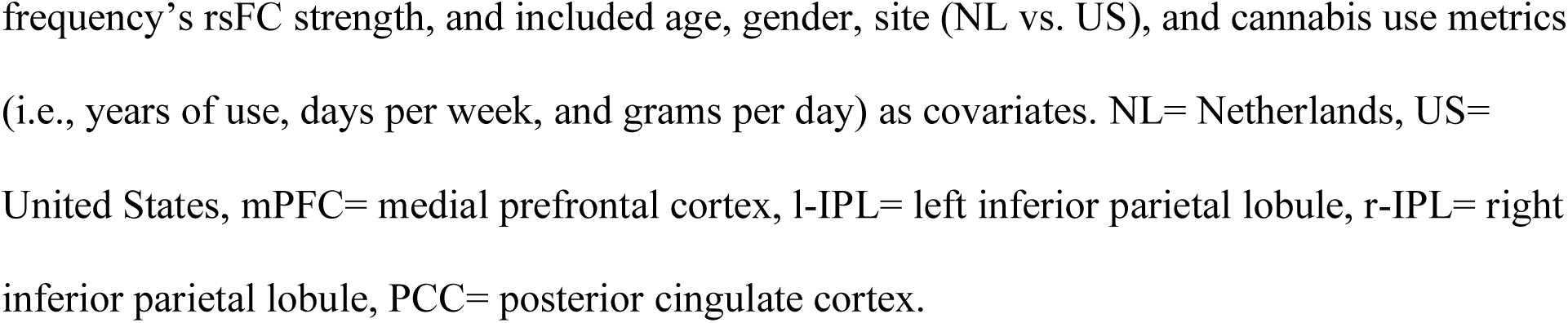
Participants by Group (n) and mean rsFC strength in Each ROI-ROI Model.

|  |  |  |  |  |
| --- | --- | --- | --- | --- |
| *ROI-ROI rsFC* | *No Pain (n)*<br>*(M/SE)* | *Sometimes (n)*<br>*(M/SE)* | *≥ Half the Time (n)*<br>*(M/SE)* | *Total (N)* |
| mPFC-l-IPL | 45<br>(-.54/.13) | 46<br>(-.92/.12) | 20<br>(-.65/.19) | 111 |
| mPFC- r-IPL | 44<br>(-.57/.1) | 47<br>(-.78/.1) | 22<br>(-.63/.15) | 113 |
| mPFC-PCC | 44<br>(-.82/.14) | 44<br>(-1.14/.14) | 21<br>(-.85/.21) | 109 |
| l-IPL-r-IPL | 47<br>(.05/.08) | 46<br>(-.14/.08) | 22<br>(-.21/.11) | 115 |
| l-IPL-PCC | 47<br>(-.24/.09) | 46<br>(-.64/.09) | 21<br>(-.51/.14) | 114 |
| r-IPL-PCC | 47<br>(-.3/.09) | 46<br>(-.7/.09) | 20<br>(-.44/.14) | 113 |

*Note.* The number of participants included in each Generalized Linear Model using likelihood ratio tests and Type III Wald $\chi^2$ statistics evaluating the main effects and the interaction terms of pain frequency and cannabis use measures on resting-state functional connectivity (rsFC) strength. One-way ANCOVA's were used to obtain the means and standard errors of each pain
frequency's rsFC strength, and included age, gender, site (NL vs. US), and cannabis use metrics (i.e., years of use, days per week, and grams per day) as covariates. NL= Netherlands, US= United States, mPFC= medial prefrontal cortex, l-IPL= left inferior parietal lobule, r-IPL= right inferior parietal lobule, PCC= posterior cingulate cortex.

**Table 3.**
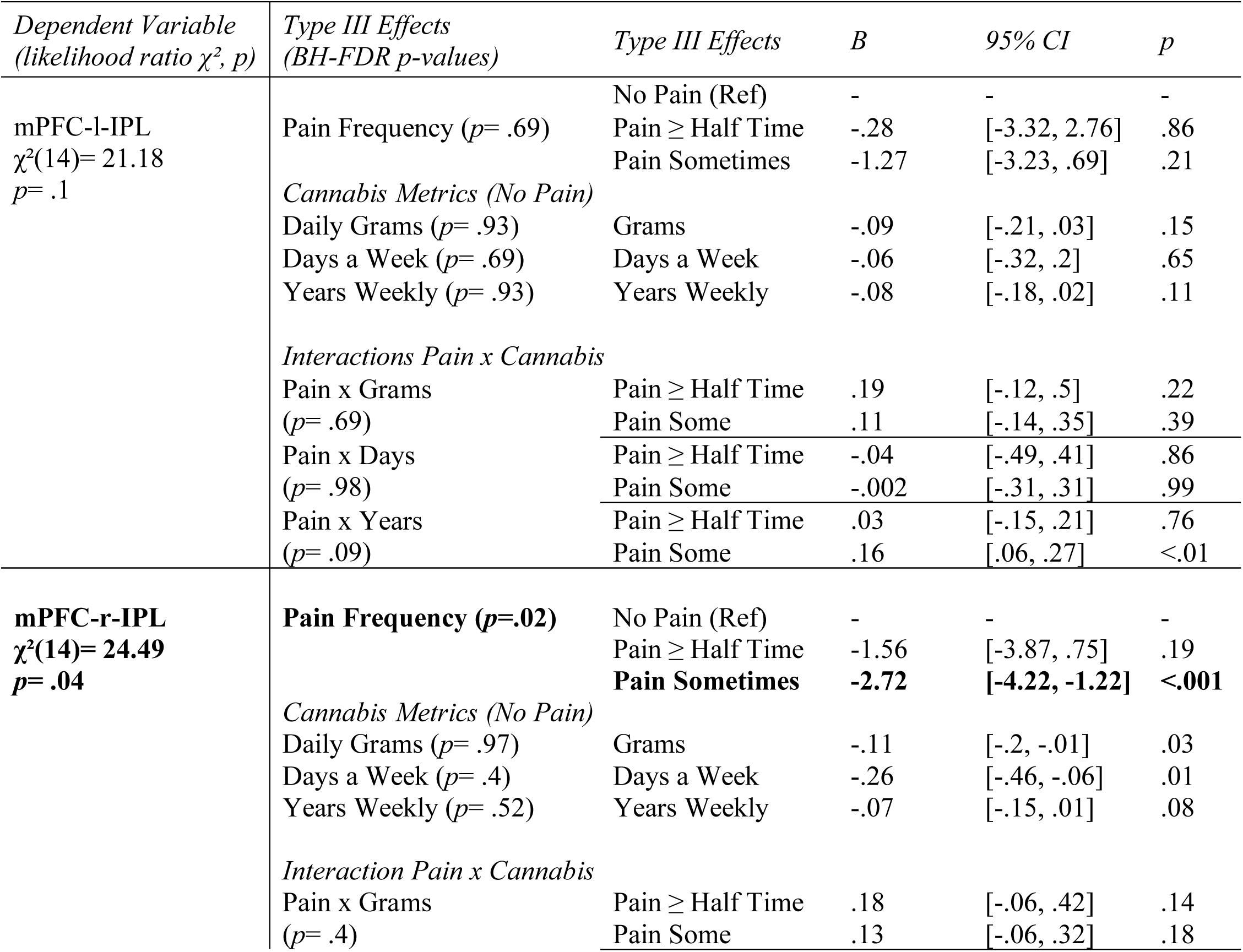
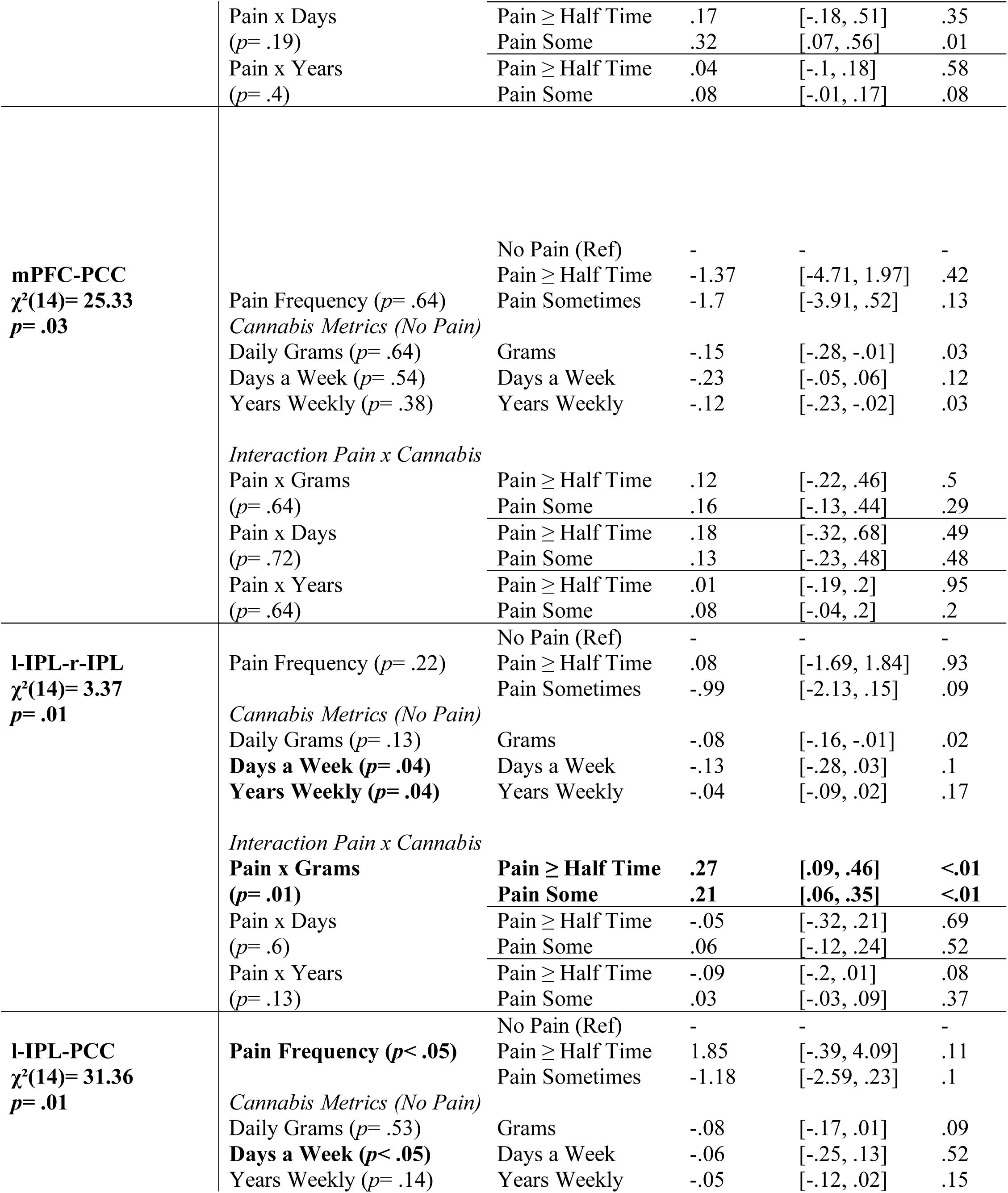
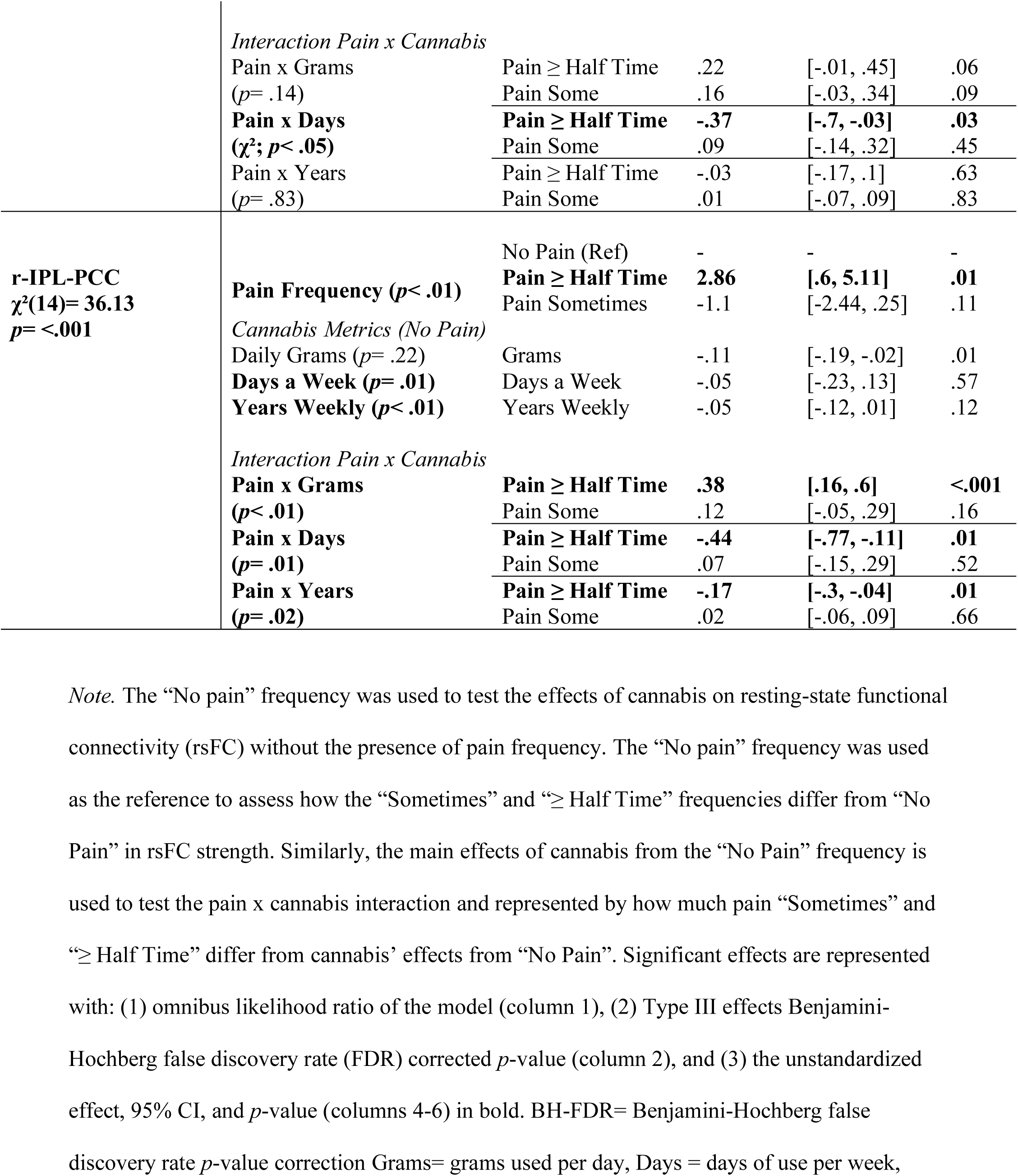
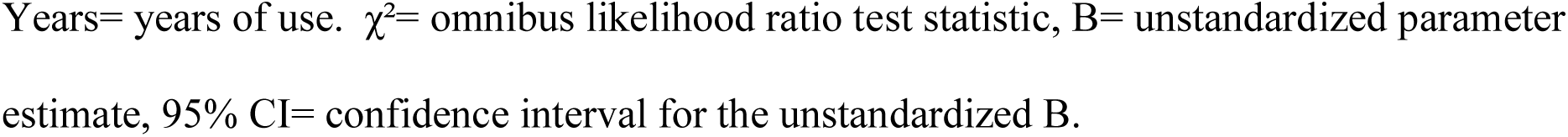
Generalized Linear Models using Pain Frequency, Cannabis Use, and their Interaction as Predictors of Region of Interest (ROI)-ROI Default Mode Network (DMN) resting-state functional connectivity (rsFC) Strength

### Main effects of pain frequency on DMN rsFC

Compared to “no pain”, increased pain frequency significantly predicted mPFC-r-IPL rsFC strength (Wald χ²(2)= 12.68, *p*< .01; Type III: BH-FDR *p*= .02), r-IPL-PCC rsFC strength (Wald χ²(2)= 14.06, *p*< .001; Type III BH-FDR *p*< .01), and l-IPL-PCC rsFC strength (Wald χ²(2)= 9.13, *p*= .01; Type III: BH-FDR *p*< .05).

mPFC-r-IPL rsFC strength was significantly weaker for pain “sometimes” (*B*= -2.72, 95% CI [-4.22, -1.22], *p*< .001), and for pain “≥ half the time” but not significant (*B*= -1.56, 95% CI [-3.87, .75], *p*= .19). Contrarily, r-IPL-PCC rsFC strength was stronger for the “≥ half the time” pain frequency (*B*= 2.86, 95% CI [.6, 5.11], *p*= .01), whereas pain “sometimes” showed weaker strength but not significant (*B*= -1.1, 95% CI [-2.44, .25], *p*= .11). Although pain frequency predicted l-IPL-PCC rsFC strength overall, there were no significant effects of pain “sometimes” (*B*= -1.18, 95% CI [-2.59, .23], *p*= .1) or pain “ ≥ half-time” (*B*= 1.85, 95% CI [- .39, 4.09], *p*= .11).

No significant effects of pain frequency were observed for mPFC-l-IPL, mPFC-PCC, or l-IPL-r-IPL rsFC (all p > .05).

### Main effects of cannabis use behaviors on DMN rsFC

The days per week of cannabis use significantly predicted l-IPL-r-IPL rsFC (Wald χ²(1)= 6.61, *p*= .01; Type III BH-FDR *p*= .04), l-IPL-PCC rsFC (Wald χ²(1)= 6.41, *p*= .01; Type III BH-FDR *p*< .05), and r-IPL-PCC rsFC (Wald χ²(1)= 8.51, *p*< .01; Type III BH-FDR *p*= .01). More days of weekly cannabis use was associated with weaker rsFC strength across these rsFC strengths but not significant (l-IPL-r-IPL: *B*= -.13, 95% CI [-.28, .03], *p*= .1; l-IPL-PCC: *B*= -.06, 95% CI [-.25, .13], *p*= .57; r-IPL-PCC: *B*= -.05, 95% CI [-.12, .13], *p*= .57).

Years of weekly use significantly predicted mPFC-PCC rsFC strength (Wald χ²(1)= 4.31, *p*= .04) with more years of weekly use associating with weaker mPFC-PCC rsFC (*B*= -.08, 95% CI [-.15, -.01], *p*= .04). However, the Type III effect did not survive BH-FDR corrections (*p*= .09). Years of weekly cannabis use also predicted l-IPL-r-IPL connectivity (Wald χ²(1)= 6.2, *p*= .01; Type III BH-FDR *p*= .04), and r-IPL-PCC connectivity (Wald χ²(1)= 12.78, *p*< .001; Type III BH-FDR *p*< .01). Longer durations of weekly cannabis use were associated with weaker rsFC strength for both, but these effects were not significant (l-IPL-r-IPL: *B*= -.04, 95% CI [-.09, .02], *p*= .17; r-IPL-PCC: *B*= -.05, 95% CI [-.12, .01], *p*= .12).

Daily grams used significantly predicted l-IPL-r-IPL rsFC strength (Wald χ²(1)= 3.93, *p*<.05), such that more grams associated with weaker rsFC strength (*B*= -.08, 95% CI [-.16, -.01], *p*< .05), although the Type III effect did not survive BH-FDR corrections (BH-FDR *p*= .1)

No other cannabis use main effects significantly predicted rsFC strength (all p > .05).

### Cannabis x pain interaction in rsFC

There were significant pain frequency x days a week interactions on l-IPL-PCC connectivity (Wald χ²(2)= 8.39, *p*= .02; Type III BH-FDR *p*< .05) and r-IPL-PCC connectivity (Wald χ²(2)= 8.67, *p*= .01; Type III BH-FDR *p*= .03). More days a week of use for pain “≥ half the time” was associated with weaker l-IPL-PCC rsFC (*B*= -.37, 95% CI [-.7, -.03], *p*= .03) and r-IPL-PCC rsFC (*B*= -.77, 95% CI [-.77, -.11], *p*= .01). However, this effect was not observed for pain “sometimes” (l-IPL-PCC: *B*= .09, 95% CI [-.14, .32], *p*= .09; r-IPL-PCC: *B*= .07, 95% CI [- .15, 29], *p*= .52) (see Figures 1 and 2).

**Figure 1.**
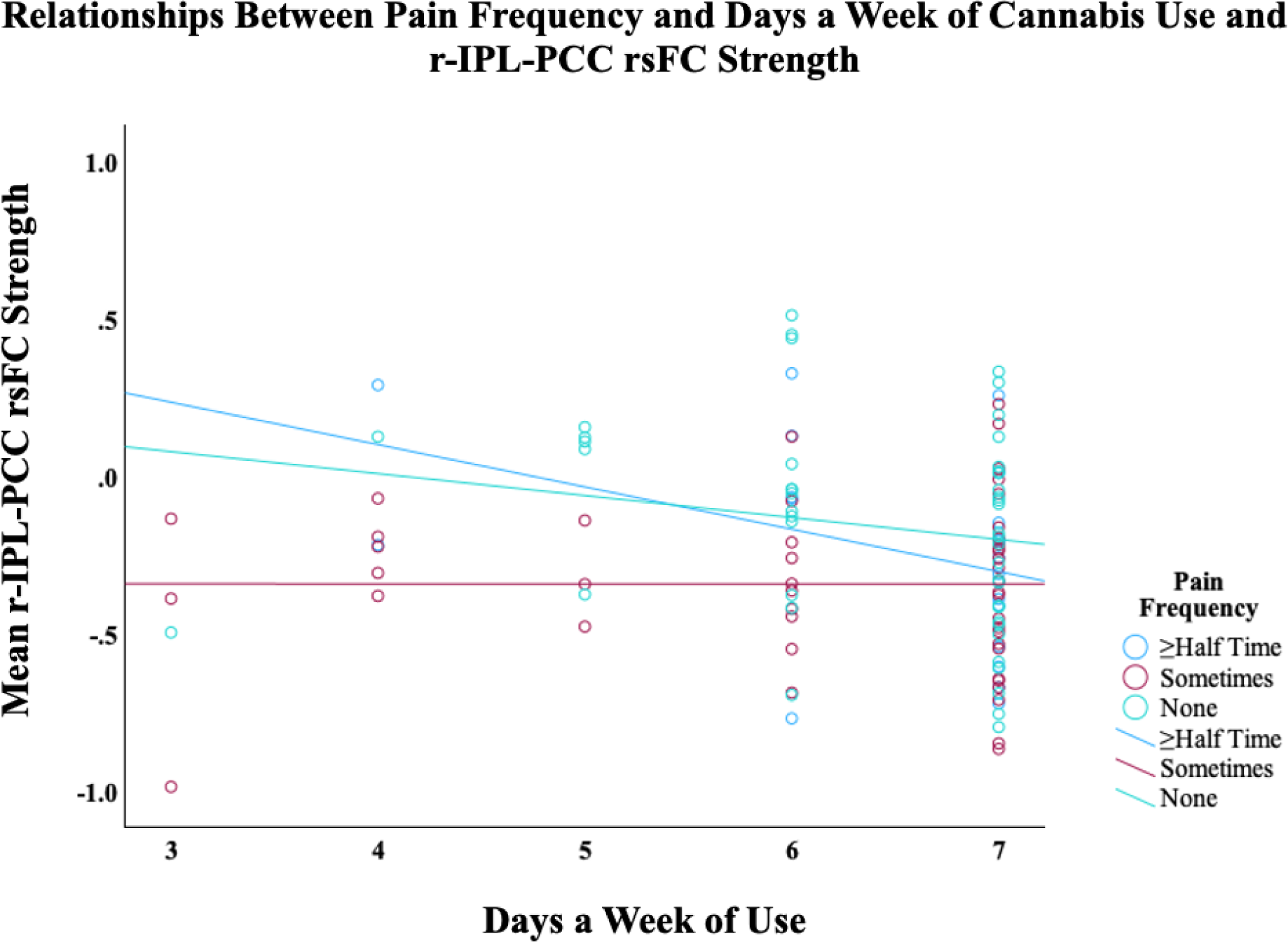
Plotted linear relationships of the effects of days of cannabis use per week on resting-state functional connectivity (rsFC) strength between the right inferior parietal lobule (r-IPL) and the posterior cingulate cortex (PCC). Those with pain “≥ half the time” had significantly weaker connectivity with more cannabis use days per week compared to “no pain”.

**Figure 2.**
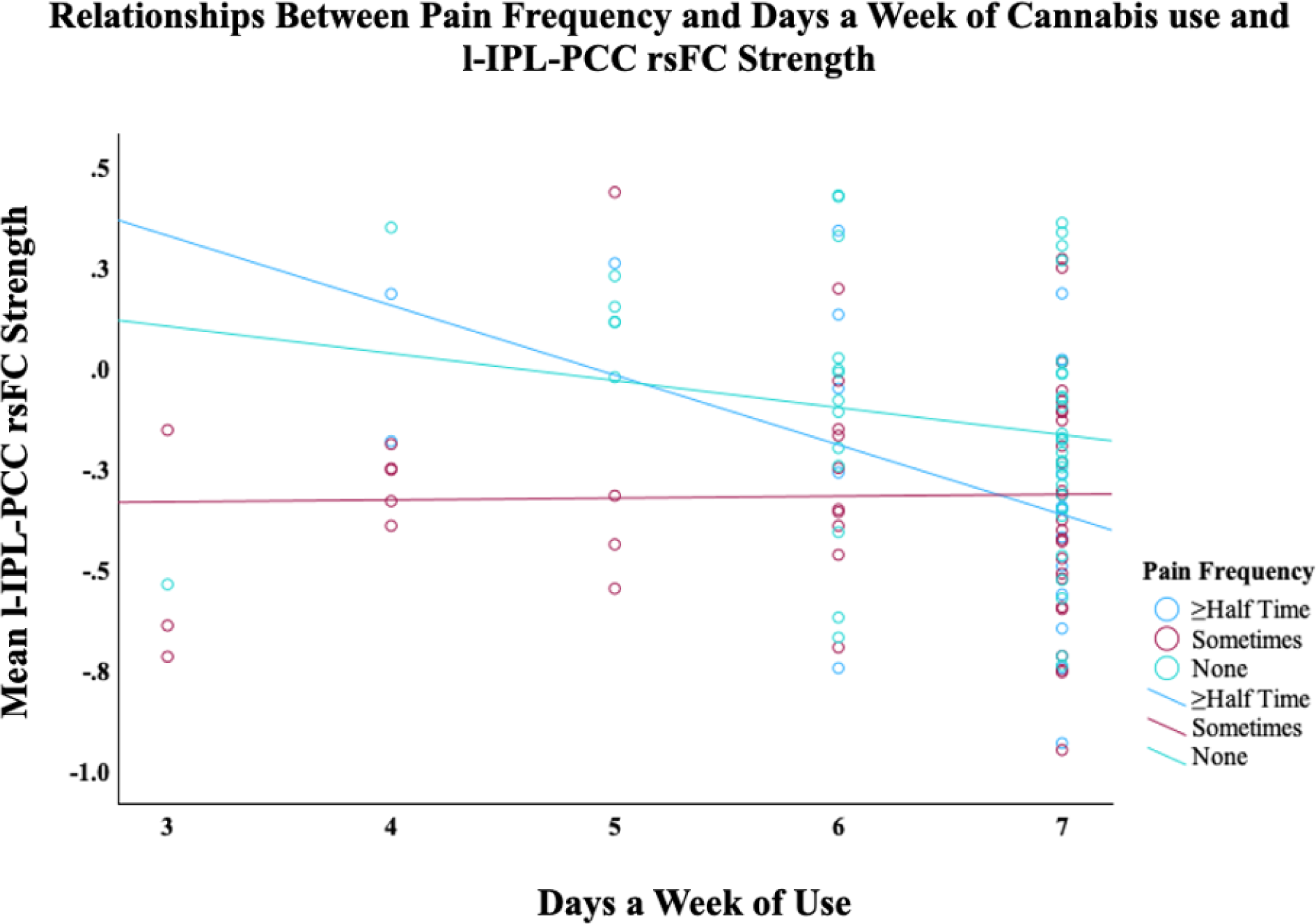
Plotted linear relationships of the effects of days of cannabis use per week on resting-state functional connectivity (rsFC) strength between the left inferior parietal lobule (l-IPL) and the posterior cingulate cortex (PCC). Those with pain “≥ half the time” had significantly weaker connectivity with more cannabis use days per week compared to “no pain”.

Similarly, the pain frequency x years of weekly use interaction on r-IPL-PCC (Wald χ²(2)= 8.67, *p*= .01; Type III BH-FDR *p*= .03) indicated that more years of weekly use reduced r-IPL-PCC rsFC for pain “≥ half the time” (*B*= -.17, 95% CI [-.3, -.04], *p*= .01), but not in pain “sometimes” (*B*= .02, 95% CI [-.06, .09], *p*= .66) (see Figure 3).

**Figure 3.**
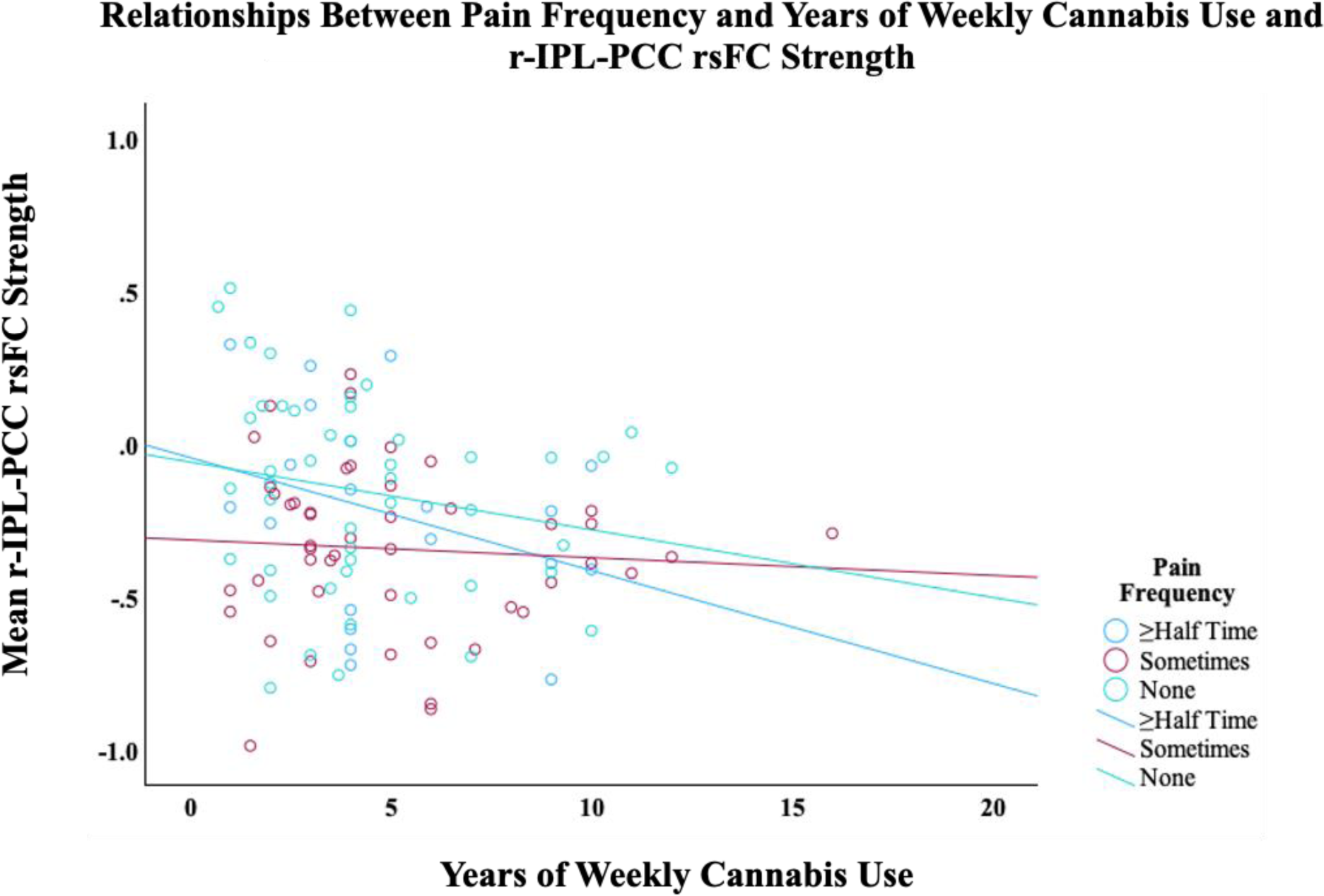
Plotted linear relationships of years of weekly cannabis use on resting-state functional connectivity (rsFC) strength between the right inferior parietal lobule (r-IPL) and the posterior cingulate cortex (PCC). Those with pain “≥ half the time” had significantly weaker connectivity with more years of weekly use compared to “no pain”.

There were significant interactions of pain frequency x daily grams on l-IPL-r-IPL rsFC (Wald χ²(2)= 13.66, *p*= .001; Type III BH-FDR *p*= .01) and r-IPL-PCC rsFC (Wald χ²(2)= 12.32, *p*< .001; Type III BH-FDR *p*< .01). Geater grams per day was associated with stronger l-IPL-r-IPL rsFC for pain “≥ half the time” and “sometimes” (≥half the time: *B*= .27, 95% CI [.09, .46], *p*< .01; sometimes: *B*= .21, 95% CI [.06, .35], *p*= .01). More grams per day associated with significantly stronger r-IPL-PCC rsFC for pain “≥ half the time” (*B*= .38, 95% CI [.16, .6], *p*< .001), but not significantly stronger for pain “sometimes” (*B*= .12, 95% CI [-.05, .29], *p*= .16) (see Figure 4).

**Figure 4.**
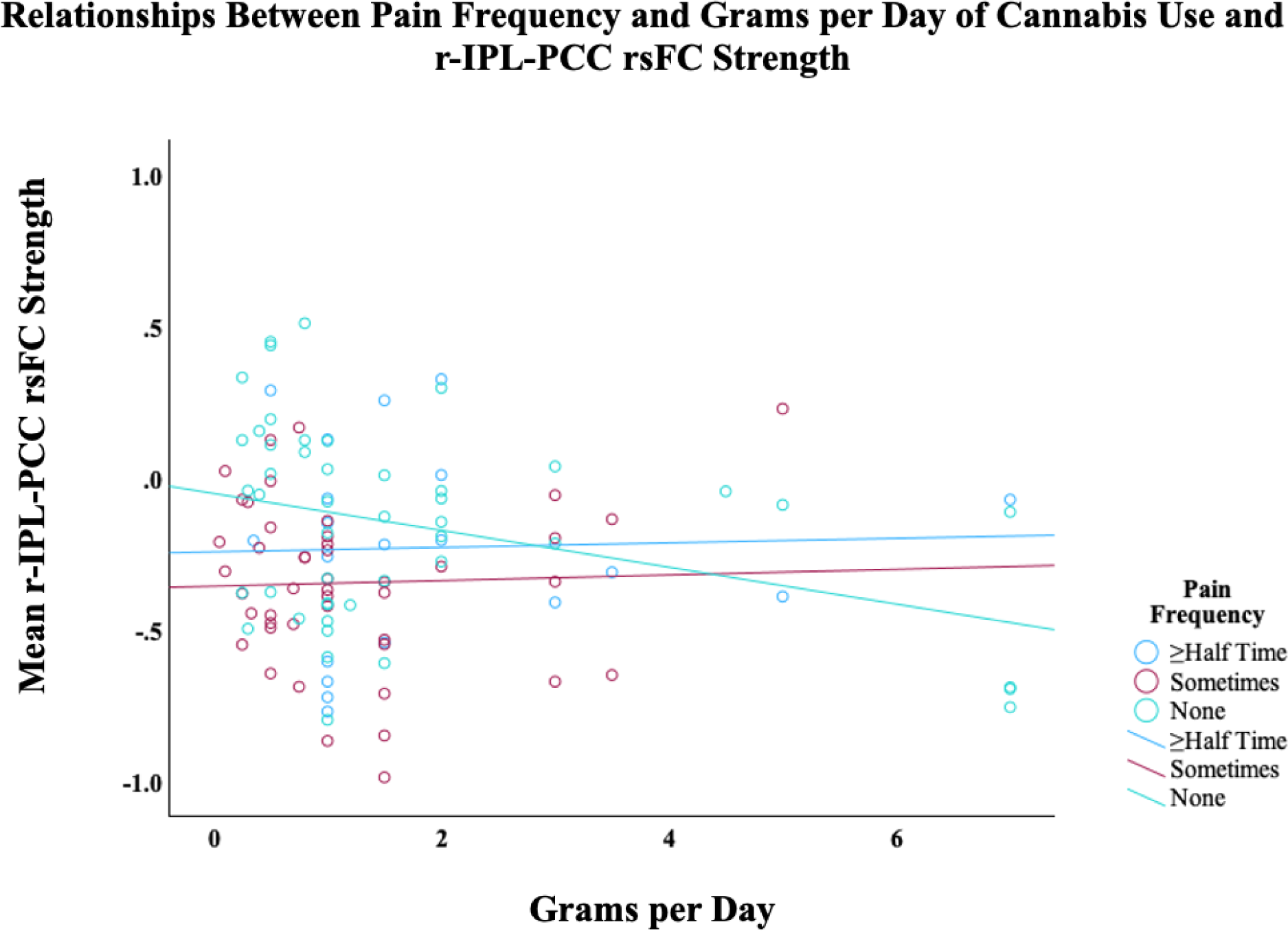
Plotted linear relationships of the effects of daily grams of cannabis used on resting-state functional connectivity (rsFC) strength between the left inferior parietal lobule (l-IPL) and the posterior cingulate cortex (PCC). Those with pain “sometimes” and “≥ half the time” had significantly stronger connectivity with more grams used compared to “no pain”.

No other significant interactions were found on the rsFC strengths.

## Discussion

We examined how pain frequency and cannabis use interact on rsFC strength within the DMN. Our findings showed that more frequent pain had a negative association with frequent as well as long-term cannabis use in IPL-PCC rsFC. On the other hand, more frequent pain had a positive association with daily consumption of cannabis in bilateral IPL rsFC. These findings suggest that in individuals with recurring pain, cannabis use is linked to dysregulated DMN coupling that may reflect altered self-referential integration and regulation of pain-related internal processing.

We found that pain frequency interacted with frequency and duration of cannabis use such that greater weekly frequency of use, as well as longer duration of use, were associated with greater reductions in bilateral IPL-PCC rsFC, reflecting a longer-term decoupling pattern linked to cumulative use in those with frequent pain. This pattern can be interpreted as reflecting longer-term alteration characterized by reduced coordination between regions involved in internally directed processing, attention, and pain-related self-monitoring. In other words, in people who experience pain more often, more sustained cannabis involvement may be linked to a degradation of communication between these DMN region. Persistent pain has been shown to alter the ECS’s signaling in corticolimbic regions that disrupt the excitation-inhibition balance (19,29). Reduced CB1 receptor availability from chronic cannabis exposure may further diminish the buffering capacity and increase susceptibility to network-level disintegration (17,18). Thus, pain-related ECS alterations and cannabis-related CB1 downregulation may result in weaker IPL-PCC rsFC, concordant with existing studies (30,31).

Persistent pain engages salience and interoceptive processing systems, which can bias the brain toward internally focused threat monitoring and reduce flexible switching between DMN and externally oriented networks (32). For example, the PCC is a central DMN hub implicated in internally directed cognition, including self-referential evaluation, integration of internal bodily states, and coordination of DMN connectivity (33). Prior work links weaker IPL-PCC rsFC with chronic pain and has been related to abnormalities in pain rumination and persistent pain-related cognition (9). Our findings of weaker IPL-PCC rsFC in the present study may be interpreted as reduced coordination between parietal integration processes from cannabis use and may reflect (1) diminished integration of cognitive control of self-referential pain representations, (2) less stable DMN internal-state integration, and/or (3) maladaptive DMN reorganization that supports repetitive internal focus on pain rather than flexible regulation (10,34,35). Thus, when these systems are altered by both frequent pain and more frequent and/or long-term cannabis use, top-down regulation of internally oriented attention to pain may be further dysregulated (9). Together, the interaction of cannabis use and pain frequency suggests that frequent pain may create a vulnerability context such that weaker IPL-PCC rsFC is moderated by greater cumulative cannabis exposure.

In contrast, greater pain frequency with higher *daily* grams of cannabis was associated with greater l-IPL-r-IPL and r-IPL-PCC connectivity. This pattern in the opposite direction may reflect a more proximal, state-like, or compensatory effect of heavier current use, as opposed to a cumulative effect as measured by frequency of *weekly* use or *total duration* of use. One possibility is that, in the context of frequent pain, larger day-level cannabis intake is linked to increased coupling in DMN as the brain adapts to ongoing nociceptive burden and acute pharmacologic effects. Existing studies demonstrate a role of IPLin both acute as well as long-term effects of pain in self-referential cognition, attentional processes (9,10,36). Acutely, Baliki et al. (2008) reported that greater engagement of posterior DMN regions during spontaneous pain fluctuations. On the other hand, Hemington et al (2016 reported increased IPL coupling with chronic pain, suggesting increased engagement of internally directed cognitive processes during ongoing pain (37).

### Limitations and Conclusions

Our results demonstrated that in the context of more frequent pain, cumulative cannabis exposure (greater weekly use and more years of use) was associated with weaker IPL-PCC rsFC, whereas heavier current use (greater daily grams) was associated with stronger l-IPL-r-IPL and r-IPL-PCC rsFC. These divergent interaction effects of pain frequency and dimensions of cannabis use suggest that, in the context of more frequent pain, cannabis use may relate to DMN rsFC through distinct neural processes that depend on cumulative vs. proximal effects.

These findings should be interpreted with some important considerations. First, this study measured pain via a single-item self-report and may be limited to detect larger effects given the smaller sample of those reporting pain “most of the time”. Future studies should include additional measures of pain, such as intensity, duration, affective impact, that a more thorough assessment of pain provides a multidimensional characterization of pain, allowing for better identification of its severity, chronicity, functional impact, and clinical relevance than a single-item question.

To conclude, these findings demonstrate that pain frequency is a key context shaping the neurobiological correlates of exposure to cannabis, highlighting a subgroup of individuals who use cannabis that may differ in risk, adaptation, and treatment needs beyond what cannabis use measures alone capture (38). Further, these findings point to two different cannabis-related processes emerging under greater pain frequency, rather than one simple linear effect.

## Data Availability

All data produced in the present study are available upon reasonable request to the authors

## Notes

### Competing Interest Statement

The authors have declared no competing interest.

### Funding Statement

Funding for this research was obtained through a grant, 1R01 DA042490, awarded to Francesca Filbey and Janna Cousijn from the National Institute on Drug Abuse/National Institute of Health.

## References

1. Cooke ME, Potter KW, Jashinski J, Pascale M, Schuster RM, Tervo-Clemmens B, et al. Development of cannabis use disorder in medical cannabis users: A 9-month follow-up of a randomized clinical trial testing effects of medical cannabis card ownership. Front Psychiatry. 2023 Mar 7;14. doi:10.3389/fpsyt.2023.1083334

2. Dawson D, Stjepanović D, Lorenzetti V, Cheung C, Hall W, Leung J. The prevalence of cannabis use disorders in people who use medicinal cannabis: A systematic review and meta-analysis. Drug Alcohol Depend. 2024 Apr;257:111263. doi:10.1016/j.drugalcdep.2024.111263

3. Duerden EG, Albanese M. Localization of pain-related brain activation: A meta-analysis of neuroimaging data. Hum Brain Mapp. 2013 Jan;34(1):109–49. doi:10.1002/hbm.21416

4. Davis KD, Moayedi M. Central Mechanisms of Pain Revealed Through Functional and Structural MRI. Journal of Neuroimmune Pharmacology. 2013 Jun 24;8(3):518–34. doi:10.1007/s11481-012-9386-8

5. Peyron R, Laurent B, García-Larrea L. Functional imaging of brain responses to pain. A review and meta-analysis (2000). Neurophysiologie Clinique/Clinical Neurophysiology. 2000 Oct;30(5):263–88. doi:10.1016/S0987-7053(00)00227-6

6. Ossipov MH, Morimura K, Porreca F. Descending pain modulation and chronification of pain. Curr Opin Support Palliat Care. 2014 Jun;8(2):143–51. doi:10.1097/SPC.0000000000000055

7. Fiúza-Fernandes J, Pereira-Mendes J, Esteves M, Radua J, Picó-Pérez M, Leite-Almeida H. Common neural correlates of chronic pain – A systematic review and meta-analysis of resting-state fMRI studies. Prog Neuropsychopharmacol Biol Psychiatry. 2025 Apr;138:111326. doi:10.1016/j.pnpbp.2025.111326

8. Apkarian AV, Baliki MN, Geha PY. Towards a theory of chronic pain. Prog Neurobiol. 2009 Feb;87(2):81–97. doi:10.1016/j.pneurobio.2008.09.018

9. Kucyi A, Davis KD. The dynamic pain connectome. Trends Neurosci. 2015 Feb;38(2):86–95. doi:10.1016/j.tins.2014.11.006

10. Baliki MN, Geha PY, Apkarian AV, Chialvo DR. Beyond Feeling: Chronic Pain Hurts the Brain, Disrupting the Default-Mode Network Dynamics. The Journal of Neuroscience. 2008 Feb 6;28(6):1398–403. doi:10.1523/JNEUROSCI.4123-07.2008

11. Jahn P, Deak B, Mayr A, Stankewitz A, Keeser D, Griffanti L, et al. Intrinsic network activity reflects the ongoing experience of chronic pain. Sci Rep. 2021 Nov 8;11(1):21870. doi:10.1038/s41598-021-01340-0

12. Jones SA, Morales AM, Holley AL, Wilson AC, Nagel BJ. Default mode network connectivity is related to pain frequency and intensity in adolescents. Neuroimage Clin. 2020;27:102326. doi:10.1016/j.nicl.2020.102326

13. Wall MB, Pope R, Freeman TP, Kowalczyk OS, Demetriou L, Mokrysz C, et al. Dissociable effects of cannabis with and without cannabidiol on the human brain’s resting-state functional connectivity. Journal of Psychopharmacology. 2019 Jul 23;33(7):822–30. doi:10.1177/0269881119841568

14. Mason NL, Theunissen EL, Hutten NRPW, Tse DHY, Toennes SW, Stiers P, et al. Cannabis induced increase in striatal glutamate associated with loss of functional corticostriatal connectivity. European Neuropsychopharmacology. 2019 Feb;29(2):247– 56. doi:10.1016/j.euroneuro.2018.12.003

15. Mason NL, Theunissen EL, Hutten NRPW, Tse DHY, Toennes SW, Jansen JFA, et al. Reduced responsiveness of the reward system is associated with tolerance to cannabis impairment in chronic users. Addiction Biology. 2021 Jan 22;26(1). doi:10.1111/adb.12870

16. Ritchay MM, Huggins AA, Wallace AL, Larson CL, Lisdahl KM. Resting state functional connectivity in the default mode network: Relationships between cannabis use, gender, and cognition in adolescents and young adults. Neuroimage Clin. 2021;30:102664. doi:10.1016/j.nicl.2021.102664

17. Hirvonen J, Goodwin RS, Li CT, Terry GE, Zoghbi SS, Morse C, et al. Reversible and regionally selective downregulation of brain cannabinoid CB1 receptors in chronic daily cannabis smokers. Mol Psychiatry. 2012 Jun 12;17(6):642–9. doi:10.1038/mp.2011.82

18. D’Souza DC, Cortes-Briones JA, Ranganathan M, Thurnauer H, Creatura G, Surti T, et al. Rapid Changes in Cannabinoid 1 Receptor Availability in Cannabis-Dependent Male Subjects After Abstinence From Cannabis. Biol Psychiatry Cogn Neurosci Neuroimaging. 2016 Jan;1(1):60–7. doi:10.1016/j.bpsc.2015.09.008

19. Katona I, Freund TF. Multiple Functions of Endocannabinoid Signaling in the Brain. Annu Rev Neurosci. 2012 Jul 21;35(1):529–58. doi:10.1146/annurev-neuro-062111-150420

20. Kroon E, Kuhns L, Cousijn J. The short-term and long-term effects of cannabis on cognition: recent advances in the field. Curr Opin Psychol. 2021 Apr;38:49–55. doi:10.1016/j.copsyc.2020.07.005

21. Kroon E, Kuhns L, Colyer-Patel K, Filbey F, Cousijn J. Working memory-related brain activity in cannabis use disorder: The role of cross-cultural differences in cannabis attitudes. Addiction Biology. 2023 Jun 5;28(6). doi:10.1111/adb.13283

22. Kroon E, Toenders YJ, Kuhns LN, Cousijn J, Filbey F. Resting state functional connectivity in dependent cannabis users: The moderating role of cannabis attitudes. Drug Alcohol Depend. 2024 Mar;256:111090. doi:10.1016/j.drugalcdep.2024.111090

23. Narrow WE, Clarke DE, Kuramoto SJ, Kraemer HC, Kupfer DJ, Greiner L, et al. DSM-5 Field Trials in the United States and Canada, Part III: Development and Reliability Testing of a Cross-Cutting Symptom Assessment for DSM-5. American Journal of Psychiatry. 2013 Jan;170(1):71–82. doi:10.1176/appi.ajp.2012.12071000

24. Sobell LC, Kwan E, Sobell MB. Reliability of a drug history questionnaire (DHQ). Addictive Behaviors. 1995 Mar;20(2):233–41. doi:10.1016/0306-4603(94)00071-9

25. Nieto-Castanon A. Functional Connectivity measures. In: Handbook of functional connectivity Magnetic Resonance Imaging methods in CONN. Hilbert Press; 2020. p. 26–62. doi:10.56441/hilbertpress.2207.6601

26. Von Korff M, DeBar LL, Krebs EE, Kerns RD, Deyo RA, Keefe FJ. Graded chronic pain scale revised: mild, bothersome, and high-impact chronic pain. Pain. 2020 Mar;161(3):651–61. doi:10.1097/j.pain.0000000000001758

27. Benjamini Y, Hochberg Y. Controlling the False Discovery Rate: A Practical and Powerful Approach to Multiple Testing. J R Stat Soc Series B Stat Methodol. 1995 Jan 1;57(1):289–300. doi:10.1111/j.2517-6161.1995.tb02031.x

28. Cousineau D, Chartier S. Outliers detection and treatment: a review. Int J Psychol Res (Medellin). 2010 Jun 30;3(1):58–67. doi:10.21500/20112084.844

29. Finn DP, Haroutounian S, Hohmann AG, Krane E, Soliman N, Rice ASC. Cannabinoids, the endocannabinoid system, and pain: a review of preclinical studies. Pain. 2021 Jul;162(1):S5–25. doi:10.1097/j.pain.0000000000002268

30. Pujol J, Blanco-Hinojo L, Batalla A, López-Solà M, Harrison BJ, Soriano-Mas C, et al. Functional connectivity alterations in brain networks relevant to self-awareness in chronic cannabis users. J Psychiatr Res. 2014 Apr;51:68–78. doi:10.1016/j.jpsychires.2013.12.008

31. Lorenzetti V, Lubman DI, Whittle S, Solowij N, Yücel M. Structural MRI Findings in Long-Term Cannabis Users: What Do We Know? Subst Use Misuse. 2010 Jun 30;45(11):1787–808. doi:10.3109/10826084.2010.482443

32. Wiech K, Ploner M, Tracey I. Neurocognitive aspects of pain perception. Trends Cogn Sci. 2008 Aug;12(8):306–13. doi:10.1016/j.tics.2008.05.005

33. Leech R, Sharp DJ. The role of the posterior cingulate cortex in cognition and disease. Brain. 2014 Jan;137(1):12–32. doi:10.1093/brain/awt162

34. Alshelh Z, Marciszewski KK, Akhter R, Di Pietro F, Mills EP, Vickers ER, et al. Disruption of default mode network dynamics in acute and chronic pain states. Neuroimage Clin. 2018;17:222–31. doi:10.1016/j.nicl.2017.10.019

35. Apkarian VA, Hashmi JA, Baliki MN. Pain and the brain: Specificity and plasticity of the brain in clinical chronic pain. Pain. 2011 Mar;152(3):S49–64. doi:10.1016/j.pain.2010.11.010

36. Kucyi A, Salomons T V., Davis KD. Mind wandering away from pain dynamically engages antinociceptive and default mode brain networks. Proceedings of the National Academy of Sciences. 2013 Nov 12;110(46):18692–7. doi:10.1073/pnas.1312902110

37. Hemington KS, Wu Q, Kucyi A, Inman RD, Davis KD. Abnormal cross-network functional connectivity in chronic pain and its association with clinical symptoms. Brain Struct Funct. 2016 Nov 15;221(8):4203–19. doi:10.1007/s00429-015-1161-1

38. Schlag AK, O’Sullivan SE, Zafar RR, Nutt DJ. Current controversies in medical cannabis: Recent developments in human clinical applications and potential therapeutics. Neuropharmacology. 2021 Jun;191:108586. doi:10.1016/j.neuropharm.2021.108586

